# A Machine-Learning Based Objective Measure for ALS Disease Severity

**DOI:** 10.1101/2022.01.21.22269641

**Authors:** Fernando G. Vieira, Subhashini Venugopalan, Alan S. Premasiri, Maeve McNally, Aren Jansen, Kevin McCloskey, Michael P. Brenner, Steven Perrin

## Abstract

Amyotrophic Lateral Sclerosis (ALS) disease severity is usually measured using the subjective, questionnaire-based revised ALS Functional Rating Scale (ALSFRS-R). An objective measure for tracking disease severity and progression would be a powerful tool for evaluating real-world drug effectiveness, efficacy in clinical trials, as well as identifying participants for cohort studies. Here we develop a machine learning (ML) based objective measure for ALS disease severity, based on voice samples and accelerometer measurements from a longitudinal dataset collected by the ALS Therapy Development Institute (ALS-TDI) over four years. 584 people living with ALS consented and carried out prescribed speaking and limb-based tasks. 542 participants contributed 5814 voice recordings, and 350 contributed 13009 accelerometer samples, while simultaneously measuring ALSFRS-R scores. Using the data from 475 participants, we trained machine learning (ML) models to predict bulbar-related FRS scores from speech, and limb-related scores from accelerometer samples. On the test set (n=109 participants) the voice models achieved a multiclass AUC of 0.86 (95% CI, 0.85-0.88) on speech FRS prediction, whereas the accelerometer models achieved a median multiclass AUC of 0.73 on 6 limb-related functions. Further, the correlation (and inverse correlations) across different bulbar and limb functions observed in the self-reported ALSFRS-R scores are preserved in the ML-derived scores. Next, we used these models and self-reported ALSFRS-R scores to evaluate the real-world effects of edaravone, a drug recently approved for use in ALS. In the cohort of 54 test participants who received edaravone as part of their usual care, the ML-derived objective scores were consistent with the self-reported ALSFRS-R scores, and both showed highly variable outcomes from edaravone treatment from person to person. At the individual level, the continuous ML-derived score is able to capture a gradual change that is absent in the integer ALSFRS-R scores. This demonstrates the value of these tools for assessing both disease severity and potentially drug effects.

## Introduction

ALS is a progressive motor neuron disease presenting with both upper and lower motor neuron signs. It is generally agreed that clinical symptoms and pathophysiology in ALS are usually focal initially, spreading contiguously from the onset site in both upper motor neuron (UMN) and lower motor neuron (LMN) compartments (Charcot, 1874; Swash, 1986; Ravits, 2007; Ravits, 2007; Sekiguchi, 2014). However, clinical presentations of motor neuron diseases and ALS are highly heterogeneous - particularly at symptom onset and early during disease (Swinnen, 2014). Hence, medical assessments of ALS onset and severity rely on subjective evaluation of overall functionality of the patient, particularly through qualitative evaluation of UMN, LMN and bulbar symptoms. However, the design of efficient clinical trials largely requires identifying signals of therapeutic efficacy in large numbers of patients with demonstrably similar rates of disease progression. This makes it challenging to test many promising therapeutics. The combination of qualitative symptom evaluation and heterogeneous disease presentation are major challenges for both disease diagnosis, prognosis, and for identifying effective therapeutics (Richards, 2020). For instance, of more than 54 potential therapeutics having been tested in ALS clinical trials, only one (riluzole) has been shown to extend survival, and only two (riluzole and edaravone) may marginally slow ALS disease progression (Lacomblez, 1996; Miller, 2012; Rothstein, 2017). Thus, the lack of diagnostic and/or prognostic biomarkers for ALS and the similar lack of objective clinical outcome measures (that balance robustness and sensitivity) have contributed to inefficient and unsuccessful efforts to effectively treat or cure ALS (Cudkowicz, 2010; Nicholson, 2015; Mitsumoto, 2015). As a step to overcome this, our work aims to provide an objective measure for ALS disease severity.

The current standard tool for monitoring ALS disease severity is the ALS Functional Rating Scale (ALSFRS) and ALS Functional Rating Scale-Revised (ALSFRS-R) (Cederbaum, 1996; Cederbaum, 1999). These measures are based on multiple-choice questionnaires designed to assess the global function of the person living with ALS. Specifically, the ALSFRS-R questionnaire asks participants to rate their functional abilities on an integer scale of 0 (“can’t do”) to 4 (“normal ability”) on three bulbar functions (speech, salivation, swallowing), six limb-related functions (handwriting, cutting_food, dressing_hygiene, turning_in_bed, walking, climbing_stairs), and three respiratory functions (dyspnea, orthopnea, respiratory_insufficiency). Individual scores are summed to produce a global score of between 0=worst and 48=best. As the disease progresses, global function measurably declines (Cederbaum, 1996; Cederbaum, 1999). ALSFRS-R is a non-invasive and cost-effective approach to monitor disease severity which strongly correlates with survival time in ALS patient populations and can serve as a linear predictor for the future rate of progression (Kaufmann, 2005; Kollewe, 2008).

Clinical settings utilize ALSFRS-R scores assessed by a variety of examiners. Most often, scores are assigned by neurologists specializing in ALS and or nurse practitioners (Cederbaum, 1999, Atassi 2014). However, in some instances, self-assessment of ALSFRS-R has been utilized for clinical research (Smith, 2017; 2011, Wicks). Differences between the examiner - neurologist, nurse, self-report - does introduce variability in the measure (Healy, 2012; Taylor, 2016). While early studies (Franchignoni, 2013) showed low interobserver reliability due to its categorical nature, more recent studies (Berry 2019) have shown high correlation (0.93) between in-clinic ALSFRS-R and smartphone self-report. Other efforts, such as the Rasch-Built Overall Amyotrophic Lateral Sclerosis Disability Scale (ROADS) (Fournier, 2020) aim to improve on the test-retest reliability and item targeting of ALSFRS-R. However, ROADS also relies on a subjective questionnaire. Nevertheless, ALSFRS-R is still the most popular and well studied measure.

Development of objective measures of disease severity is a critical unmet need. Recent efforts (Rutkove 2018, Berry 2019) have explored frequent data collection at home using relatively inexpensive technologies including hand-grip dynamometry, electric impedance myography, speech recordings, and self-reported ALSFRS-R scores towards developing more objective ALS symptom progression endpoints. Early on, Pan 2013 used accelerometry data to study Parkinsons disease progression. Recently, Rutkove 2018 has shown that good response rates for at-home data collection is possible with good design. Similarly, Berry 2019 has shown high correlation with in-clinic and smartphone self-report of ALSFRS-R (at baseline). Our work takes these a step further.

We develop ML-based objective measures of ALS disease severity based on voice samples of prescribed speech and accelerometer measurements of limb-based tasks. Here we collected these physiological measurements - voice recordings and accelerometer recordings - together with the self-reported ALSFRS-R scores. We then build ML models, a voice model that takes as input the speech data and accelerometer models that take as input the accelerometer measurements. These models objectively assess ALS severity by learning to predict the ALSFRS-R scores corresponding to one or more functions. These were then used to compare the objective ML-predicted scores with self-reported scores to study the real-world effects of edaravone in people living with ALS.

## Data

A key requirement for developing an objective measure is collecting a dataset from a sufficiently large cohort of ALS patients. In 2014, ALS-TDI launched the Precision Medicine Program (PMP), which has enrolled more than 600 people with ALS (as of January 2021). For each participant, the PMP collects a rich dataset including biological samples (skin biopsy, whole genome sequencing and blood-based biomarkers), as well as regular measurements of self-reported ALSFRS-R scores together with physiological indicators–voice recordings and accelerometer measurements tracking prescribed limb exercises.

The dataset used in this work is derived from 584 people living with ALS, who consented to participate in the research study and contributed voice recordings, accelerometer measurements or both over about four years (Sep’14 - Aug’19). We used this data to build ML models by associating the data to self reported ALSFRS-R scores, within 60 days of their recording i.e. for each voice or accelerometer sample we associate the ALSFRS-R score that is closest to and within 60 days of the recorded sample, failing which the sample is discarded. The average time delta between recording and ALSFRS-R assessment was 3.2 days for voice and 5.3 days for accelerometer samples. This gave us 542 participants with 5,814 voice samples, and 350 participants with 13,009 accelerometer samples. To assess clinical intervention outcomes, we took advantage of the fact that a subset of the Precision Medicine Program participants had enrolled in the translational research program before the FDA approval of edaravone (Radacava). All of the participants who began using edaravone following its approval in May, 2017 (n=54) were identified^1^ and placed in the test set. The remaining participants were split randomly into the train, validation and test set to get an overall ratio of 70:15:15. Demographic details and distribution of the participants (and recordings) in the splits for model development are reported in Table 1.

**Table 1.** Demographic and clinical characteristics of participants including the number of participants and number of recordings/measurements (in parenthesis) in the train, validation, and test sets. The rightmost column shows the set of participants in the Edaravone cohort (a subset of the test set).

| Model | Train | Validation | Test | Edaravone cohort<br>(subset of Test) |
| --- | --- | --- | --- | --- |
| # total participants | 395 | 80 | 109 | 54 |
| # voice participants | 389 | 63 | 90 | 49 |
| # voice recordings | 3776 | 705 | 1333 | 832 |
| # accelerometer participants | 209 | 58 | 83 | 44 |
| # accelerometer measurements | 7448 | 2028 | 3533 | 2061 |
| Age in years (standard deviation) | 58.69 (11.79) | 57.83 (12.15) | 59.35 (10.48) | 59.41 (10.59) |
| Sex (Male/Female) | 261 / 134 | 52 / 28 | 66 / 43 | 33 / 21 |
| Median days in program (Q1, Q3) | 322 (114, 770.5) | 465.5 (170.5, 880.5) | 541 (303, 945) | 742.5 (493, 1059) |
| Avg. ALSFRS-R scores at enrollment (standard deviation) | 38.20 (7.32) | 39.89 (7.05) | 39.32 (6.78) | 40.69 (4.54) |
| Avg. ALSFRS-R scores at final point (standard deviation) | 30.25 (11.56) | 30.74 (12.11) | 28.94 (12.59) | 28.85 (10.36) |
| Onset-type (Limb / Bulbar / Respiratory / Asymptomatic) | 311 / 59 / 22 / 3 | 67 / 7 / 5 / 1 | 86 / 19 / 2 / 2 | 45 / 9 / 0 / 0 |

The speech data consisted of audio recordings (collected via phone) of participants speaking a sentence, “I owe you a yoyo today” (Green, 2013), repeated five times. The participants login to a secure portal and opt-in to receive an automated call where they utter the sentence that gets recorded. Most participants have multiple recordings taken every few weeks over a year or more.

The accelerometer measurements, obtained from Actigraph GT3X devices (one for each limb), came from 5 limb-based exercises each about 45s long with a short 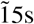 break in between. A full set of measurements is approximately 5 minutes in length (Supplementary Material-Video of movements).

## Results

### Approach Overview

Using the physiological measurements - voice and accelerometer recordings - and the self-reported ALSFRS-R scores, we developed 2 types of models: (1) voice model and (2) accelerometer model. The voice model is a convolutional neural network (CNN) that takes as input the speech recording and was trained to predict the probability distribution over the 5 ALSFRS-R score classes (0-4) corresponding to the speech (bulbar) function. This was trained on 3776 speech samples from 389 participants. The set of accelerometer models take as input the accelerometer measurements and were trained to predict the probability distribution over the 5 ALSFRS-R score classes (0-4) for 9 functions (the 6 limb-related and 3 respiratory functions). We developed different accelerometer models to compare the performance of (a) various architectures - CNN, linear regression, logistic regression, MLP), (b) input types - high resolution (Raw 30Hz) or down-sampled versions (Uniform-1Hz and FFT-1Hz) and (c) output types - a single function’s score or all 9 functions’ score jointly (multi-label). These were trained on 7448 accelerometer measurements from 209 participants. The details of the data processing and the models themselves are described in the Methods section. Our results in this section report the performance of the single best performing accelerometer model (MLP multi-label FFT-1Hz) and the voice CNN model.

## Analysis

### ALSFRS-R analyses

ALSFRS-R data of 109 test participants were analyzed for pairwise correlation between individual ALSFRS-R assessments (Figure 1). Strong correlations were found (a) between speech, salivation, and swallowing (*R*^2^ = 0.74-0.80), (b) between handwriting, cutting food, and dressing/hygiene (*R*^2^ = 0.69-0.80), and (c) between scores meant to evaluate lower limb functions (walking and climbing stairs) (*R*^2^ = 0.8). Unsurprisingly, dyspnea, orthopnea, and respiratory insufficiency self-assessments were also strongly correlated (*R*^2^ 0.74-0.76) (Figure 1A). Interestingly, some ALSFRS-R metrics of progression were very poorly correlated. For example, scores for respiratory functions were not correlated with scores for bulbar functions, and neither were scores for limb functions correlated with scores for bulbar functions, e.g., scores for walking were also not correlated with scores for salivation or speech (*R*^2^ = 0.02 and 0.08) (Figure 1A).

**Figure 1.**
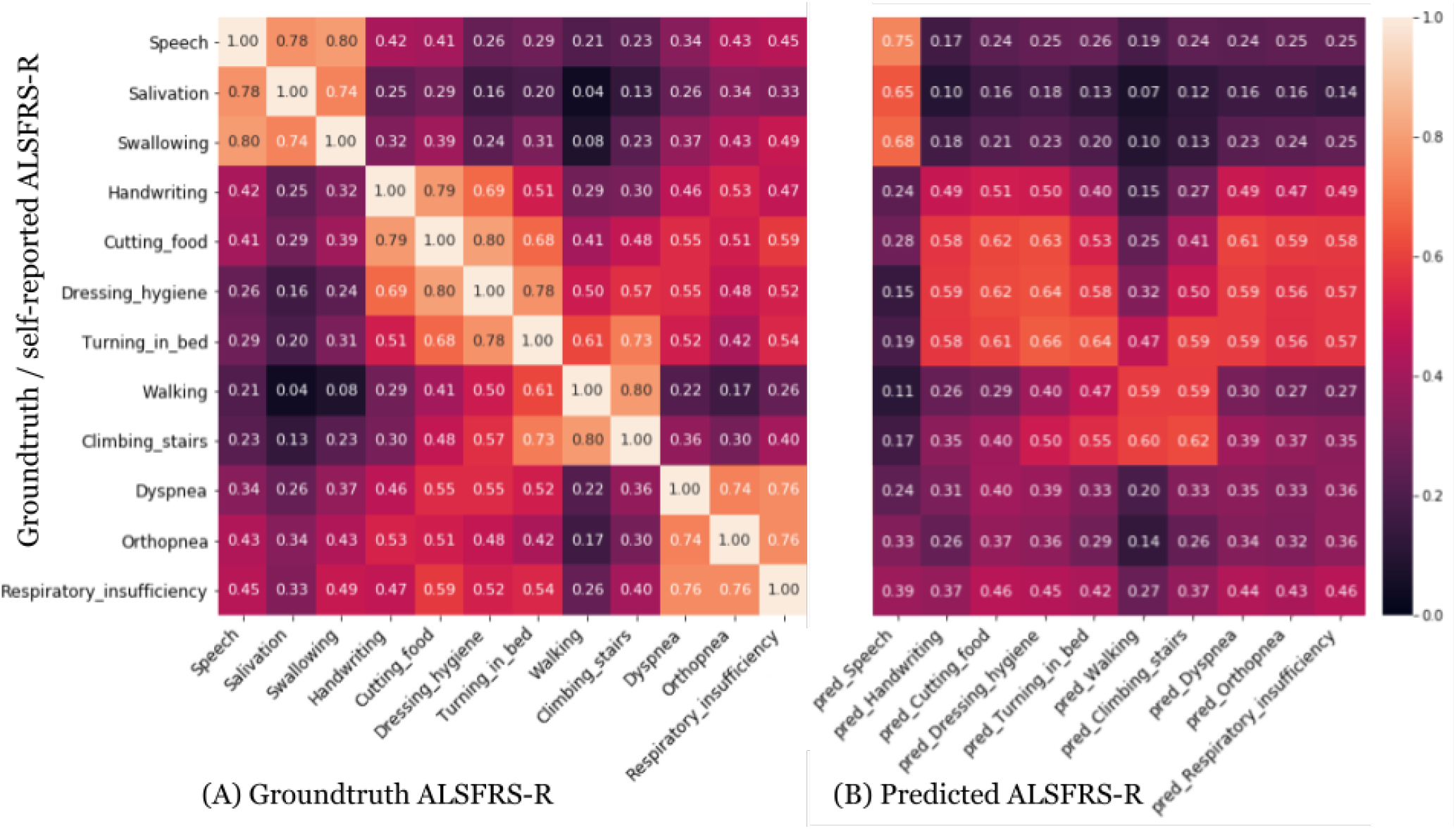
Correlation between ground-truth (y-axis) ALSFRS-R scores with (A) Groundtruth ALSFRS-R and (B) ML-predicted ALSFRS-R scores (x-axis) on the full test set of 109 participants. (A) There is a strong correlation between ALSFRS-R scores for speech, salivation, and swallowing. We also observed correlations between other sets of limb-related functions, specifically, handwriting, cutting food, dressing hygiene and turning in bed, and then between walking, and climbing stairs, and the respiratory-related functions dyspnea, orthopnea, and respiratory insufficiency. (B) We observed that the predicted ALSFRS-R for speech (based on the voice model) is most correlated with groundtruth speech ALSFRS-R followed by strong correlations with salivation and swallowing. Additionally, as with the ground-truth FRS scores, we observed that the accelerometer models’ predictions for the limb-related functions (handwriting, cutting_food, dressing_hygiene, turning_in_bed, walking, climbing_stairs) are also correlated. We further note that the accelerometer models’ predictions and the speech models are not correlated with the respiratory-related functions’ scores.

### ML-predicted ALSFRS-R vs. Ground-truth self-reported ALSFRS-R

#### Correlation between ground-truth and Predicted ALSFRS-R scores for all functions

We defined self-reported ALSFRS-R values as ground-truth against which to compare predicted ALSFRS-R values derived from objective data collection tools, including voice recordings or accelerometer recordings. The ML-predicted ALSFRS-R scores for speech derived from voice recordings were strongly correlated with ground-truth scores for speech, salivation, and swallowing (*R*^2^ = 0.75, 0.65, and 0.68 respectively) (Figure 1B). Similarly, accelerometer reading-derived ALSFRS-R score predictions for handwriting, cutting food, dressing/hygiene, turning in bed, walking, and climbing stairs were correlated with their respective ground-truth ALSFRS-R values (*R*^2^ = 0.49, 0.62, 0.64, 0.64, 0.59, and 0.62 respectively). Unsurprisingly, neither voice recording derived ALSFRS-R speech predictions, nor accelerometer-based ALSFRS-R limb predictions achieved high correlation with respiratory function ALSFRS-R ground-truth scores, with the highest *R*^2^ at 0.59 (Figure 1B).

#### Correlations at baseline and correlation of slopes

Next we slice the correlations by looking at the participants’ scores at baseline (when they enrolled into the PMP) and their slopes over time. Figure 2 presents the correlation between ground-truth and model predicted ALSFRS-R scores at baseline for speech (Figure 2A) and the average of 6 limb functions (Figure 2C). The correlation for speech scores at baseline is 0.80 and that of the limb functions at baseline is 0.67. The correlation of the slopes for the speech scores is 0.68 (Figure 2B), and is 0.60 for the slopes from the limb scores (Figure 2D). Not surprisingly, the correlation of the slopes computed over time from the ground-truth and predicted values is lower for both models compared to the correlations of the raw scores at just the baseline which is just a single point in time.

**Figure 2.**
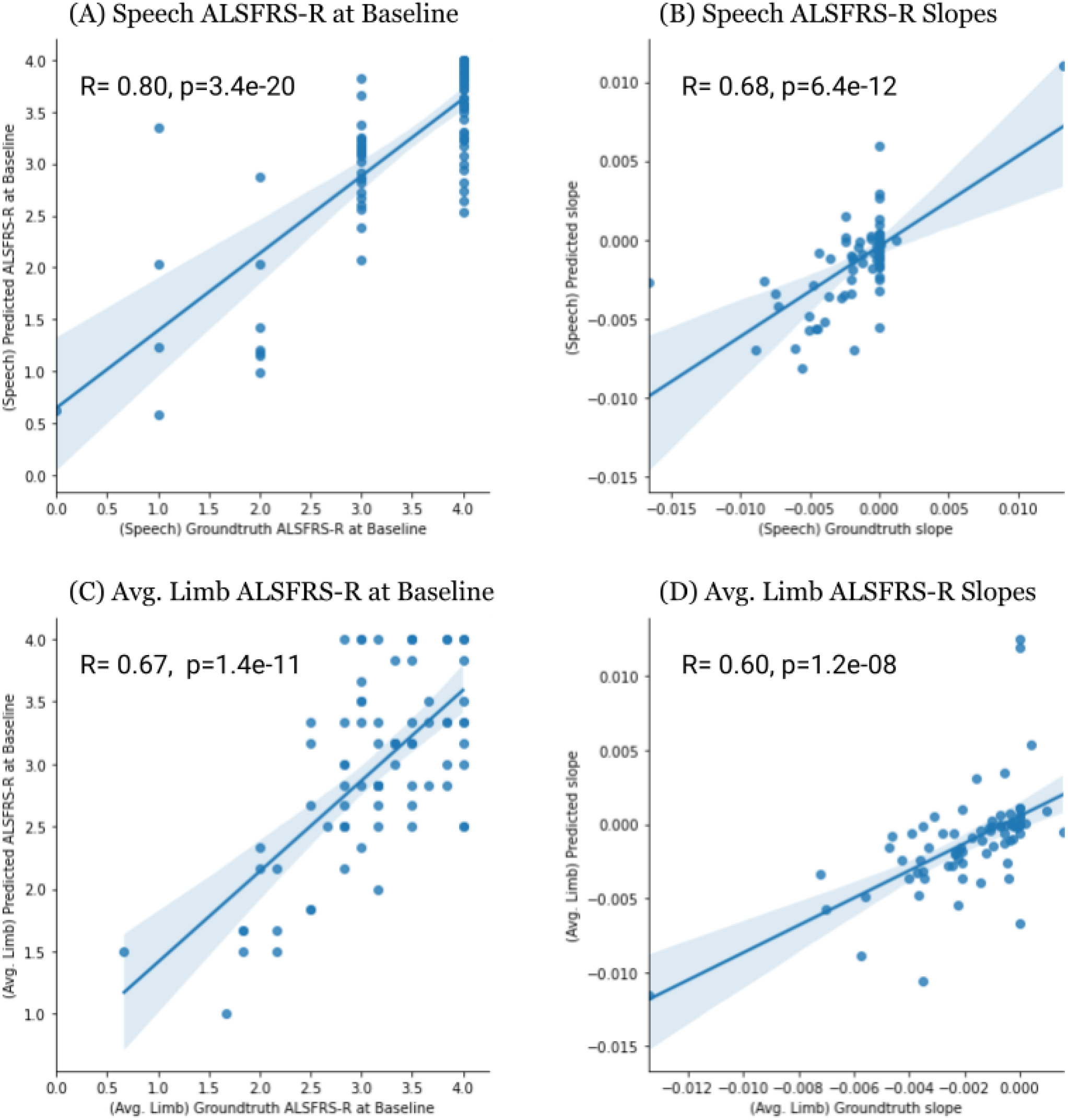
Plots depicting correlation of ALSFRS-R scores at baseline (PMP enrollment) and correlation of slopes over time on the 109 test participants. The plots depict pearson correlation between ground-truth and predicted scores based on (A) speech ALSFRS-R scores at baseline, (B) slopes over time from ALSFRS-R speech scores, (C) limb ALSFRS-R scores (averaged over 6 limb-based movements) at baseline, and (D) slopes over time from averaged limb-movement scores. The x-axis in the subfigures denotes the ground-truth values, and the y-axis denotes the predicted values. Subfigures A and B for speech include scores on 90 participants, subfigures C and D for limb show scores on 83 participants.

#### ROC-AUC values for the predicted scores

Table 2 presents ROC-AUC values with bootstrapped confidence intervals for the predicted ALSFRS-R scores for speech, 6 limb-related functions, and the 3 respiratory functions for our best performing models, using the voice CNN model and accelerometer (FFT 1Hz MLP multi-label) model. Confusion matrices for these two models’ predictions are presented in Supplementary Table 1a and 1b. The voice CNN model, evaluated on 1333 samples from 90 test participants, attained the highest AUC (0.86 [CI:0.85-0.88]). For the limb related functions, the MLP multi-label FFT-1Hz model evaluated on 3833 accelerometer samples from 83 test participants performed best, with an AUC between 0.70 - 0.75 on 5 of the 6 limb related functions, with the lowest performance for handwriting (AUC = 0.64). Somewhat surprisingly, the accelerometer model also achieved AUCs between 0.63 - 0.75 on the respiratory functions. Supplementary Table 2 presents performance of other accelerometer model variants, and compares it with the previous best method of normalizing the accelerometer data based on TBVM (Premasiri et al, 2017). Prediction of the models on individual participant recordings on 54 test participants in the edaravone cohort, along with self-reported ALSFRS-R speech scores and slopes, are shown in Supplementary Figure 3 (voice) and Supplementary Figure 4 (accelerometer).

**Table 2.** AUC values for the predicted ALSFRS-R scores for speech, and 6 limb-related functions using the voice (small CNN) model and accelerometer (FFT 1Hz MLP multi-label) model described in the Methods section. Confidence intervals (95% CI) were calculated by bootstrapping. Results for other models are reported in Supplementary Table 2.

| Function | AUC | 95%CI | Source Model |
| --- | --- | --- | --- |
| frs_speech | 0.865 | [0.847 - 0.884] | Speech |
| frs_climbing_stairs | 0.701 | [0.691 - 0.712] | Accelerometer |
| frs_cutting_food | 0.733 | [0.723 - 0.743] | Accelerometer |
| frs_dressing_hygiene | 0.729 | [0.719 - 0.742] | Accelerometer |
| frs_handwriting | 0.645 | [0.634 - 0.658] | Accelerometer |
| frs_turning_in_bed | 0.755 | [0.745 - 0.766] | Accelerometer |
| frs_walking | 0.756 | [0.746 - 0.766] | Accelerometer |
| frs_dyspnea | 0.652 | [0.636 - 0.666] | Accelerometer |
| frs_orthopnea | 0.723 | [0.706 - 0.740] | Accelerometer |
| frs_respiratory_insufficiency | 0.736 | [0.721 - 0.752] | Accelerometer |

### Changes in ML-predicted and self-reported ALSFRS-R scores with real-world Edaravone use in PMP Participants with ALS

Having established that voice and accelerometer recording based ALSFRS-R predictions correlate with their related ground-truth ALSFRS-R scores, we applied these technologies to study the real-world performance of edaravone, approved in the United States for the treatment of ALS disease progression, retrospectively on 54 test participants. We used the date of edaravone commencement to define time ‘0’ and plotted self-reported ALSFRS-R scores (Figure 3A and 3C), ML model predicted speech ALSFRS-R (Figure 3B), and ML model predicted limb-function ALSFRS-R (Figure 3D). Individual participant plots are in Supplementary Figures 3 and 4. Based on these figures we can observe that for an individual function (speech in particular), the ML-predicted continuous score is able to show gradual changes compared to the integer self-reported score. This is also more apparent in individual participant plots.

**Figure 3.**
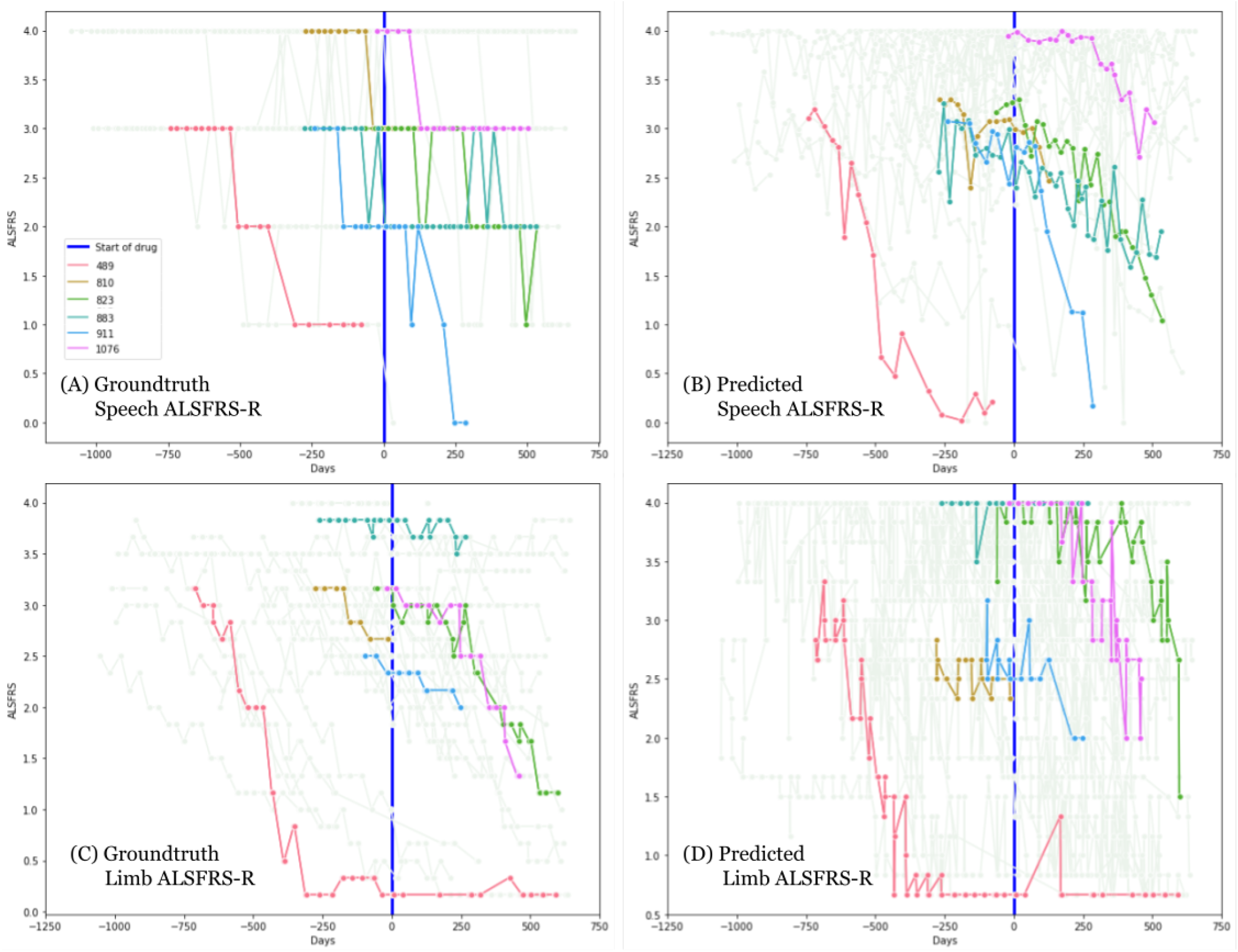
Plots depicting progression of ALS on the subset of 54 test participants in the Edaravone study. The plots depict ALSFRS-R scores based on (A) groundtruth speech scores, (B) speech scores as predicted by the voice model, (C) groundtruth limb scores (averaged over 6 limb-based movements), and (D) averaged limb-movement scores as predicted by the accelerometer model. The x-axis represents days with 0 (vertical blue line) as the point where the participant starts the drug. Y-axis represents the ALSFRS-R score. Subfigures A and B for speech include scores on 49 participants, subfigures C and D for limb show scores on 44 participants. 6 representative participants (IDs: 489, 810, 823, 883, 911, and 1076) are highlighted in color in all panels, other participants are shown in lighter color in the background. Predicted scores are plotted for each available recording (on the recorded date/day). The groundtruth and predicted scores for each of the 54 participants is included in Supplementary Fig. 3 (for voice) and Supplementary Fig. 4 (for limb).

Where there were at least 3 data points both before and after initiation of edaravone treatment, we calculated slopes (r-values) using linear regression analyses of ground-truth self-reported ALSFRS-R, predicted voice ALSFRS-R, and predicted limb-function ALSFRS-R both pre- and post-edaravone initiation. These are shown in Supplementary Figure 1A-D. Based on (Takahashi et al. 2017), to study the association of edaravone with respiratory functions, in Supplementary Figure 2A-E, we also plotted the slopes (r-values) of participants computed after initiation of edaravone against their respiratory ALSFRS-R (averaged over the 3 respiratory functions) prior to and at the time of initiation of edaravone. This compares progression of participants (slopes) computed on ground-truth respiratory, speech, and limb scores as well as the ML-predicted speech and limb scores. From these figures, we did not observe any cohort-level changes after edaravone initiation.

## Discussion

This real-world study of ALS symptom progression using digital measures - accelerometer readings and speech recordings - demonstrated the feasibility of applying digital outcomes in conjunction with machine learning (ML) to predict self-assessed ALSFRS-R. Our results, in particular, the AUC values and ALSFRS-R correlation analysis show that the predictions of the ML models are consistent with self-reported ALSFRS-R. Additionally, the continuous ML-predicted score is also able to capture the gradual transition in ALSFRS-R scores (over the duration of the study) in comparison to the integer self-reported ones (Figure 3 and Supplementary Figures 3 and 4), making it a useful tool to monitor disease severity objectively. In terms of their practical application within the PMP cohort, for the analysis of real-world use of edaravone retrospectively, both ML-predicted and self-reported ALSFRS-R did not show observable cohort-level changes (Figure 3 and Supplementary Figures 1 and 2) and indicated variable outcomes from person to person.

We were able to apply similar ML approaches to two distinct types of data: voice recordings, and limb movement as measured by accelerometry. While neither the digital voice recording phrase selected nor accelerometer-based prescribed movements were optimized to maximize signal (Allison 2019), we learned that the ML methods were still effective at predicting ALSFRS-R subscores (Table 2 and Figure 1). The effectiveness of the accelerometer based models (median multiclass AUC of 0.73 on 6 limb-related and 3 respiratory functions) was somewhat surprising because cutting food, dressing-hygiene, and handwriting can be considered largely fine-motor functions (Bakker et al, 2017), while the prescribed exercise movements captured by the accelerometer emphasized gross motor function by way of deltoid and quadriceps strength and endurance while de-emphasizing fine motor coordination. Perhaps the respiratory functions are also correlated with strength and endurance. Thus it is interesting that the ML models were still able to effectively predict functional assessment of skills that require both strength and fine motor coordination.

Another key observation from this study is how closely the predicted ALSFRS-R scores tracked variations in self-assessed ALSFRS-R scores in many cases (Figure 2, Figure 3, and Supplementary Figures 3 and 4). The correlation between ground-truth and predicted scores at baseline (Figure 2) are substantially better than the correlations of the slopes themselves. Self-assessment based measures present the risk of being subjective and, thus have low interobserver reliability (Linton 2015), and show volatility over time (Bakker et al, 2017). We had hypothesized that this volatility would not be evident using objective digital outcome measures coupled with ML tools. The volatility does manifest as lower correlation of slopes over time (Figure 2) perhaps not to the extent one would assume. Thus, although the predictions (AUCs and individual correlations) especially for speech are fairly good, there is still variance that is not captured.

As the supplementary figures show, in some instances, a participant’s self-assessed score at the time of study enrollment (baseline) was quite different from the predicted score. However, surprisingly when transient downturns or upturns in self-assessed ALSFRS-R scores were apparent, the ML/digital outcomes would sometimes vary similarly (Supplementary Figure 4). This indicates that the participants are self-consistent, in the sense that if they reported a lower score their functional ability does truly decline. This is what we believe is reflected in the ML model predictions. Although the participant might score themselves differently from a clinician, they seem to consistently associate their functional ability to the same score and are attuned to their own declines. Hence the ML model picks up on the signal. Further, since we always only consider self-assessments, this eliminates any issues due to interobserver reliability in collection of ALSFRS-R scores.

As a proof of concept, these tools were applied to the assessment of the effectiveness of edaravone in a real-world clinical setting. Neither self-assessed ALSFRS-R nor the ML/digital predictions revealed cohort-level changes in slope (Supplementary Figure 1) that might have suggested overall slowing of ALS disease progression. We also explored (in Supplementary Figure 2) whether participants who began edaravone treatment with higher respiratory function ALSFRS-R self-assessment would perform better on the drug based on the clinical trial reports demonstrating that people with ALS in Japan with higher slow vital capacity (SVC) responded to edaravone treatment while others did not (Takahashi et al. 2017). In the retrospective study, on our small cohort we did not see indications that the patients with higher self-assessed respiratory function ALSFRS-R scores performed differently than the rest of the test cohort. This requires additional study.

Important questions remain for ALS research and clinical communities. First, regarding bulbar symptom assessment, could recordings of other phrases be used to improve bulbar symptom assessment? Speech pathologists with experience in ALS assessment have been developing batteries of phrases for improved bulbar symptom assessment (Green 2018). Deploying part or all of these batteries and coupling them with ML tools could reveal even more sensitive disease progression measures. Second, we used a limited set of prescribed movements to capture arm and leg function using accelerometers. The prescribed movements have not been optimized to capture fine motor function or gross coordination. More work needs to be done to improve these protocols. Third, the PMP did not include any direct or surrogate sensory measures of respiratory function that could be used in developing ML models. That is worth further investigation.

Overall, this work demonstrates the value of digital outcome measures, specifically voice recordings and accelerometry, to study ALS. It shows that ML can be applied to such digital outcome measures to objectively predict ALS disease severity and to monitor and reveal progression patterns.. Further, the proposed measure was used to assess edaravone’s real-world performance retrospectively, on a small cohort of people with ALS enrolled in the ALS-TDI PMP, as a proof of concept demonstration. This is the first study to combine digital clinical outcome measures with ML to study an approved medication’s post-market effectiveness for ALS. Our work suggests that the proposed methods can be helpful in assessing medicines used to treat ALS, without imposing additional financial or travel burden on patients. Such an approach may be amenable for use in clinical trials, but may also be an essential strategy for assessing experimental therapies made available outside of the clinical trial setting in expanded access programs.

## Methods

This research program has been conducted in accordance with the ethical principles posited in the Declaration of Helsinki - Ethical Principles for Medical Research Involving Human Subjects. Protocol approval was provided by the institutional review board (ADVARRA CIRBI).

### Data Preprocessing

The speech data was uniformly resampled to 8kHz mono-stream. The audio samples are then converted to spectrograms, the details of which are described in the voice model section. For the exercise-based accelerometer recordings, the original accelerometer measurements (on 3 axes) are recorded at 30Hz (referred to as Raw 30Hz). We also obtained a low-resolution 1Hz version from the Actigraph software [ActiLife version 6.13.3] (referred to as Raw 1Hz). We used measurements from 4 exercises, one from each of the four limbs, i.e. Left Ankle (LA), Left Wrist (LW), Right Ankle (RA), and Right Wrist (RW). A fifth exercise involving both wrists together, which was less emphasized, was often missing and thus discarded. For all our models, we considered measurements from the four limbs, and built and evaluated models on the 30Hz data or the following variants derived from the 1Hz data:

- Total Body Vector Magnitude (*TBVM*): We reproduced this baseline from (Premasiri et al. 2017) which is based on the Raw 1Hz measurements where each limb value is normalized. To normalize, a control 1Hz vector magnitude (VM) dataset was created by collecting four to six months of accelerometer data from 18 healthy volunteers. They calculated the average VM from each limb across the prescribed movements from the healthy volunteer cohort and chose the largest value to create vector magnitude normalization coefficient for each prescribed movement (i.e., they divide by the largest value to get a scaling coefficient for each limb in the health volunteer cohort, and multiply by that co-efficient to normalize data from participants; we report these values in the Supplementary materials). Following this process (Premasiri et al, 2017), for each month of VM data for each patient, we normalized each prescribed movement VM and summarized the VM for all limbs into a single value by adding them together to obtain the TBVM value.
- *FFT 1Hz*: We applied a discrete Fast Fourier Transform on the Raw 1Hz data and for each of the 4 limbs, leading to a total of 8 features for each accelerometer measurement.
- *Uniform 1Hz*: This is a variant of the Raw 1Hz data consisting of 70 measurements per limb, (truncating shorter samples or padding zeros to longer samples). We normalized the training set’s values and applied the same parameters to normalize the validation and test sets.

### Voice Model

To build a model for predicting ALSFRS-R scores from the voice recordings, we used a convolutional neural network (CNN) architecture proposed in (Hershey et al. ICASSP’17), suited for audio classification tasks. Our model and approach is illustrated in Figure 4. The details regarding selection of parameters (mentioned below) for modeling the data and the training details are described in the Supplement.

**Figure 4.**
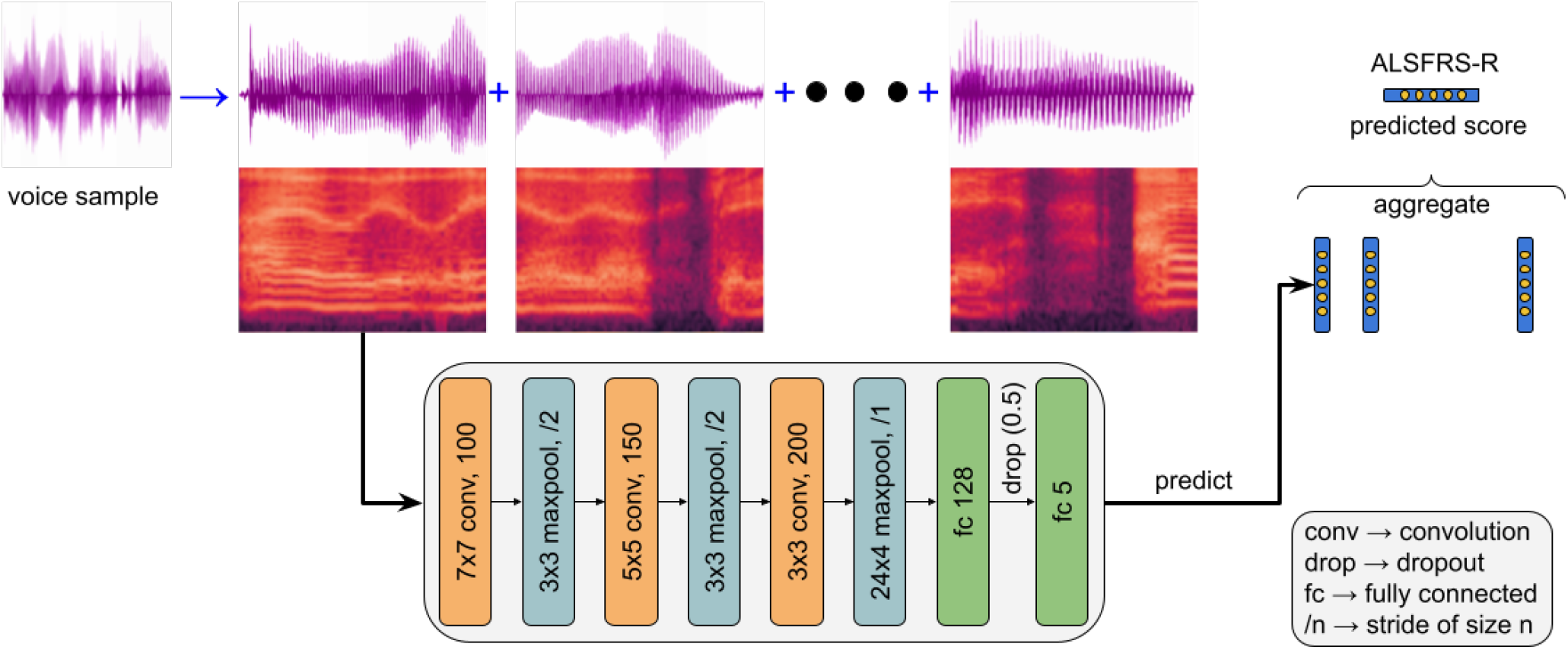
[Model overview] Our approach segments audio/accelerometer recordings into non-overlapping frames. We then convert the waveforms to derive spectrogram grayscale images. A classifier is trained on the image patches to predict the score label for each non-overlapping frame which is then aggregated to predict an ALSFRS-R score for the entire voice sample. The CNN depicted here is the architecture that was used to train the speech ALSFRS-R prediction model. (The kernel shape (mxn), and number of filters are denoted for convolutions, while for maxpool the shape and stride are noted). We note that this overall approach is also identical to how the CNN model is applied to the accelerometer data.

The model takes as input the spectrogram of the waveform, a visual representation of the spectrum of a signal’s frequencies as it varies with time. To create the spectrogram inputs, the audio recording is processed into non-overlapping 960ms audio “frames”. These frames are then decomposed with a short-time Fourier transform applied to 25ms windows every 10ms, with the last window zero-padded. The resulting spectrogram is integrated into 64 mel-spaced frequency bins, and the magnitude of each bin is log-transformed after adding a small offset of 0.001 to avoid numerical issues. This gives log-mel spectrogram context-windows of 96×64 bins that form the input. The spectrogram windows are then input to a 2-dimensional convolutional network (architecture depicted in Figure 4) with a logistic loss for each score class (0-4). During training, all audio frames (equivalently, spectrogram context-windows) from a recording get the same ALSFRS-R score as the entire audio clip’s label, i.e., the patient’s reported speech ALSFRS-R score on the date closest to the recording. The model’s output is a probability distribution for each of the 5 ALSFRS-R score classes (0-4) for each audio frame. During training, we fetch mini-batches of 64 input examples by randomly sampling context-windows from all audio samples. Since the number and length of audio samples from each participant (and correspondingly the frequency of occurrence of the label/score) can be different, the loss for each window is weighted inversely to the frequency of the class label at the frame-level. During inference, we first aggregate the window/frame-level scores by taking a mean across all spectrogram context windows to get a probability distribution of the scores over the entire audio recording. The final ALSFRS-R score predicted by our approach is the average of the score values weighted by the model’s predicted probability of each score for the audio recording.

### Accelerometer Models

We evaluated several methods to model the accelerometer measurements. In particular, on the Uniform 1Hz data and the FFT 1Hz data, we applied linear regression, logistic regression, as well as multi-layer perceptron (MLP) models. The models are trained to predict the probability distribution over the 5 ALSFRS-R score classes (0-4) for 9 functions (the 6 limb-related and 3 respiratory functions). For each ML model (linear regression, logistic regression, MLP) and data type (Uniform 1Hz and FFT 1Hz), 9 models were trained to predict each of the 9 functions individually. Additionally, for the MLP, a tenth model that jointly predicts scores for all 9 functions was trained. This joint model is termed the multi-label classification model.

To model the high-resolution (Raw 30Hz) accelerometer data, we use a small CNN similar to the voice model described above. In this case, the accelerometer measurements are processed into non-overlapping 75s “frames.” These are decomposed with a short-time Fourier transform applied to 7s windows every 3s, with the last two windows zero-padded. These result in linear spectrogram patches of 19×129 bins. The details of these parameter choices and computation of spectrogram patch sizes are described in the Supplement. The accelerometer CNN model also differs in that it uses a multi-task classification to learn and predict scores for all the respiratory and limb-related ALSFRS-R scores. The rest of the training and evaluation procedure is identical as with the voice model.

### Evaluation metric

As described in the section on Voice model, our models output a probability for each score class (0-4) for each function, from which we derive the predicted ALSFRS-R score for each function (speech, walking etc.). To evaluate the overall performance of the model, we use the probabilities predicted for each score class to compute the ROC-AUC (1 vs all AUC) and take the average to report the multiclass AUC for each function. For all the other analysis, such as to compute correlations, or participant slopes, we directly use the predicted ALSFRS-R score for the function.

## Data Availability

All data produced in the present study are available upon reasonable request to the authors

https://github.com/pmphelp/paper-code

## Data Availability

The datasets generated and/or analyzed during the current study are available from the corresponding author on reasonable request.

## Acknowledgements

This research could not be possible without the immense contributions from people living with ALS who have participated for months and years in the ADVARRA CIRBI IRB approved Translational Research Program for ALS. We thank each participant for their contributions. We thank Oltiana Mosko, Amy Lummen, Taylor Charbonneau, Melissa Nollstadt, and Beth Levine for their stewardship of patient participants. We thank the ALS patient and caregiver community, in particular Augie’s Quest for a Cure, for financially supporting this research. We thank Philip Nelson and Rif A. Saurous from Google Research for their support and encouragement through the duration of this project, and thank Dave Parrish, Joel Shor and Dotan Emmanuel for early research contributions. We also thank Yun Liu, Akinori Mitani, Michael McConnell, and Michael Howell from Google Health for feedback on a draft of this work.

## Author Contributions Statement

S.P. and F.G.V. conceived of the translational research program that resulted in the data collected and analyzed in this manuscript. F.G.V., S.V., M.P.B., and S.P. conceived of the study objectives. S.V., M.P.B., A.J., and K.M. conceived and executed model tranining. F.G.V., A.S.P., MM, and SP contributed to data collection efforts. F.G.V., S.V., A.S.P., M.M., A.J., K.M., M.P.B., and S.P. contributed to the interpretation of the results. F.G.V. and S.V. took the lead in writing the manuscript. All authors provided critical feedback and helped shape the research, analysis, and manuscript.

## Supplementary Materials

### Exercise movements video

For the accelerometer recordings, the set of movements/exercises prescribed to participants is included as a video (approximately 5 minutes in duration) with the supplementary materials. (Supplementary Material-Video of movements)

### Predicted vs. Groundtruth ALSFRS-R Confusion Matrices

Supplementary Table 1A shows the confusion matrix of the voice ALSFRS-R values using the voice-based model predicting the speech FRS score. The model has an overall accuracy of 73% (AUC of 0.86). On recordings, with ALSFRS-R scores of 4 (normal), the model predicts them accurately on 620 of the samples’ 692 (90%) samples. On samples with scores 3 and 2, the model predicts them with an accuracy of 57% (213 of 315, and 80 of 141 respectively). The model overestimates a rating of 3 and predicts those as a 4 (29% cases), and in the case of a 2, it confuses it as a 3 or 1 in 23% and 14% of the cases, respectively. It predicts 40% of the recordings rated as 1 accurately and tends to confuse them mostly as a 0 (29%) and at other times as a 2 or a 4.It predicts 65% of the recordings with a score of 0 as 0, and the remaining as 1. In all cases, the model’s misclassifications appear reasonable since the range of variation is plus/minus 1.

Supplementary Table 1B shows the confusion matrix of the voice ALSFRS-R values using the accelerometer model. It presents the average predictions (rounded to the nearest integer) across ALSFRS-R scores pertaining to the six limb related functions (handwriting, cutting_food, dressing_hygiene, turning_in_bed, walking, climbing_stairs). As we can observe, the accelerometer model tends to overestimate the FRS score, providing a higher score if the participants rated themselves as 0,1,2 or 3. (i.e., it predicts these as 1,2,3 or 4 respectively just as often as it predicts the actual value). Overall, in 76% of the cases, the accelerometer predicts the same score or a score higher by 1 point. Performance of different accelerometer models Supplementary Table 2 shows the performance (AUC values) of all the accelerometer model variants described in the methods section on predicting the ALSFRS-R scores on the limb related functions.

### Performance on each of the test participants in the edaravone study

Supplementary Figures 3 and 4 present the groundtruth and predicted labels for each of the 54 participants in the subset of test participants in the Edaravone study. Supplementary Figure 3 shows the groundtruth speech ALSFRS-R values and predictions from the voice model, it also presents the slopes before and after (i.e. pre- and post-) beginning edaravone treatment. Supplementary Figure 4 is an analog of Supplementary Figure 3 on the accelerometer model. It presents the sum of groundtruth ALSFRS-R values on the six limb-related functions and predictions from the accelerometer model along with the slopes before and after beginning edaravone treatment.

### Normalization coefficient values used in TBVM computation

To compute TBVM values in our work, we used the process described in (Premasiri et al, 2017). To normalize, they created a control 1Hz vector magnitude (VM) dataset by collecting four to six months of accelerometer data from 18 healthy volunteers. They calculated the average VM from each limb across the prescribed movements from the healthy volunteer cohort and chose the largest value (which corresponded to the left-wrist and was 717.9) to create vector magnitude normalization coefficient for each prescribed movement (i.e. they divide by the largest value to get a scaling coefficient for each limb in the health volunteer cohort, and multiply by that co-efficient to normalize data from participants). The coefficients they obtained (and which we used after dividing the VM values by 717.9) are: 2.497844 (for the left-ankle), 2.492674(for the right-ankle), 1.0 (for the left-wrist), 1.01044 (for the right-wrist), and 3.123886 (for both wrists together).

### Voice and Accelerometer model parameter choices and training details

Developing the voice and accelerometer models from the original data samples involve a number of choices, both in terms of parameters for processing the data, as well as hyper-parameters chosen for the optimization/training process. To determine a number of these parameters, we first used a subset of the data (from Sep’14 - July’18) and created a training, validation, and test split (each containing roughly a third of the participants). This dataset was used to select many of the parameters used to model and process the data. Specifically, the duration of the spectrogram frames (960ms for audio, 300s for accelerometer), the overlap between frames (non-overlapping frames, or an overlap of 75s duration) and verify the choice of spectrogram i.e. log-mel spectrogram of 64 bins for audio, and a linear spectrogram for the accelerometer. In the case of the accelerometer frames, the spectrogram images are of size 19×129 calculated from the window-size, stride and frequency as: 75/(7-3) = 19; and 129 = 128+1 (30hz*7 rounded up to the nearest power of 2, then divided by 2, and then add 1). We treated both CNN models as a multi-class multi-label task to be able to predict ALSFRS-R scores for multiple functions. So we used a sigmoid function and a logistic loss for each label i.e. function+rating (e.g. speech-FRS-4, speech-FRS-3, and so on). With these parameters set, we used the validation set of the final dataset splits described in the ‘Data’ section to choose the training hyper-parameters.

Regarding training, our CNN voice and accelerometer models used batch normalization. We compared mini-batch sizes of 32, 64, and 128 frames, and learning rates of 1e-5, 1e-6, and 3e-6, and the Adam optimizer. While these parameters in themselves didn’t result in significant differences in performance some models trained faster achieving better performance sooner. For our final models, the voice model used a mini-batch size of 64 and learning rate of 1e-5, and the accelerometer model used a mini-batch of size 32 and a learning rate of 1e-5. The models were trained for around 25 epochs. The simpler accelerometer model variants (linear regression, logistic regression, and multi-layer perceptron) used a batch size of 100 and were trained for 60 epochs, and the model performing best on the validation set was used to run evaluations on the final test set.

## Supplementary Tables

**Supplementary Table 1 A.** Voice prediction model confusion matrix indicating the number of predictions that replicate groundtruth self-reportedl ALSFRS-R scores for speech.

| Speech | Predicted Scores |  |  |  |  |  |  |
| --- | --- | --- | --- | --- | --- | --- | --- |
|  |  | 0 | 1 | 2 | 3 | 4 | Total |
| Ground-truth scores | 0 | 13 | 7 | 0 | 0 | 0 | 20 |
|  | 1 | 29 | 41 | 14 | 5 | 14 | 103 |
|  | 2 | 5 | 20 | 80 | 33 | 3 | 141 |
|  | 3 | 6 | 12 | 35 | 213 | 110 | 376 |
|  | 4 | 8 | 0 | 6 | 58 | 620 | 692 |

**Supplementary Table 1 B.**
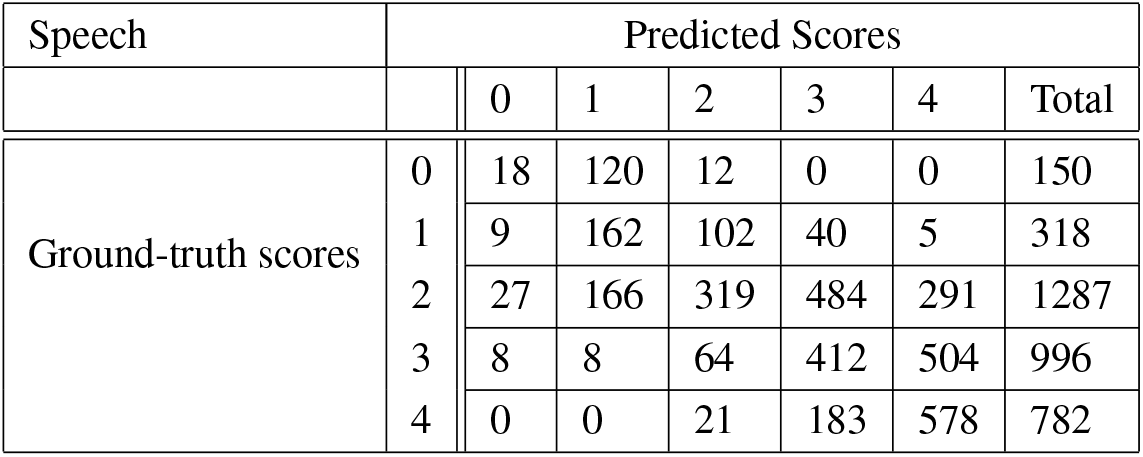
FFT Multilabel MLP model. Confusion matrix of ground-truth ALSFRS-R scores with predicted ALSFRS-R scores on the sum of the scores from the six limb related functions (handwriting, cutting_food, dressing_hygiene, turning_in_bed, walking, climbing_stairs) [Sum of limb score predictions (argmax over the sum of the limb frs prediction distributions), Groundtruth is average of the sum of limb scores and rounded to the nearest integer.]

| Speech | Predicted Scores |  |  |  |  |  |  |
| --- | --- | --- | --- | --- | --- | --- | --- |
|  |  | 0 | 1 | 2 | 3 | 4 | Total |
| Ground-truth scores | 0 | 18 | 120 | 12 | 0 | 0 | 150 |
|  | 1 | 9 | 162 | 102 | 40 | 5 | 318 |
|  | 2 | 27 | 166 | 319 | 484 | 291 | 1287 |
|  | 3 | 8 | 8 | 64 | 412 | 504 | 996 |
|  | 4 | 0 | 0 | 21 | 183 | 578 | 782 |

**Supplementary Table 2.** FFT Multilabel MLP model. Confusion matrix of ground-truth ALSFRS-R scores with predicted ALSFRS-R scores on the sum of the scores from the six limb related functions (handwriting, cutting_food, dressing_hygiene, turning_in_bed, walking, climbing_stairs) [Sum of limb score predictions (argmax over the sum of the limb frs prediction distributions), Groundtruth is average of the sum of limb scores and rounded to the nearest integer.]

| Accelerometer Data | Model | climbing_stairs | cutting_food | handwriting | turning_in_bed | walking |
| --- | --- | --- | --- | --- | --- | --- |
| TBVM | Logistic Reg. | 0.627 | 0.564 | 0.508 | 0.617 | 0.564 |
| Raw 30Hz | small CNN | 0.70 | 0.68 | 0.60 | 0.77 | 0.75 |
| Uniform 1Hz | Logistic Reg. | 0.634 | 0.593 | 0.537 | 0.602 | 0.586 |
| Uniform 1Hz | MLP (multi-label) | 0.738 | 0.726 | 0.634 | 0.753 | 0.755 |
| FFT 1Hz | MLP (multi-label) | 0.701 | 0.733 | 0.645 | 0.756 | 0.756 |

## Supplementary Figures

**Supplementary Figure 1.**
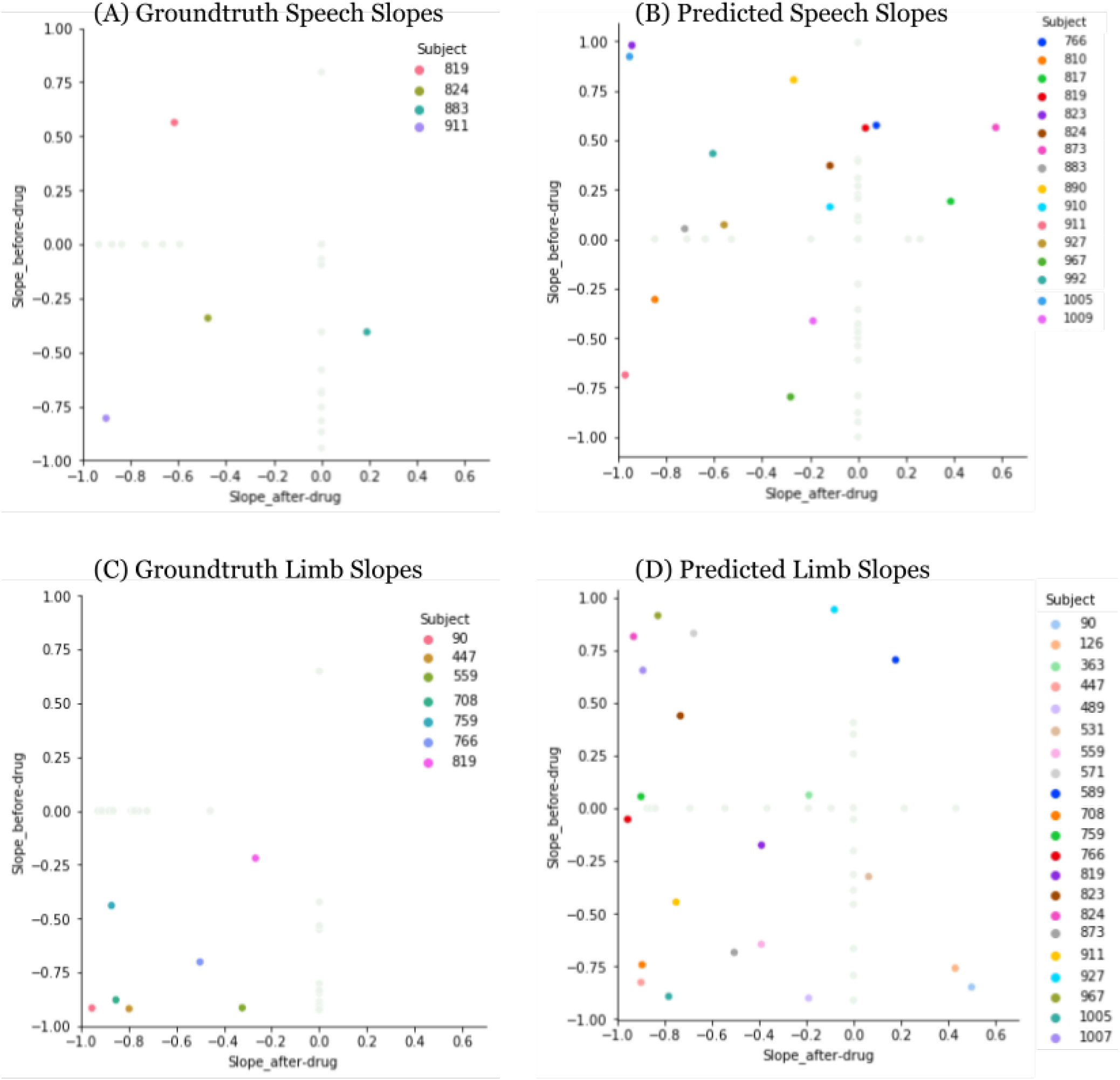
Plots comparing slopes (r-values) on the subset of 54 test participants in the Edaravone study. The plots depict slopes (r-values) computed before (pre-Edaravone) and after (post-Edaravone) starting drug using the ALSFRS-R scores: (A) slopes from groundtruth speech scores, (B) slopes from speech scores as predicted by the voice model, (C) slopes from groundtruth limb scores [scores averaged over 6 limb-based movements], and (D) slopes from averaged limb-movement scores as predicted by the accelerometer model. The x-axis represents the slope (r-value) from scores after-drug (post-Edaravone), and the y-axis represents the slope (r-value) from scores before-drug (pre-Edaravone). In each plot, the IDs of participants (subjects) who had sufficient data to compute a slope both before and after starting the drug are highlighted in color. The participants who only had sufficient data to compute slope either before or after drug are represented in a lighter color in the background (their slope is 0 in the other case).

**Supplementary Figure 2.**
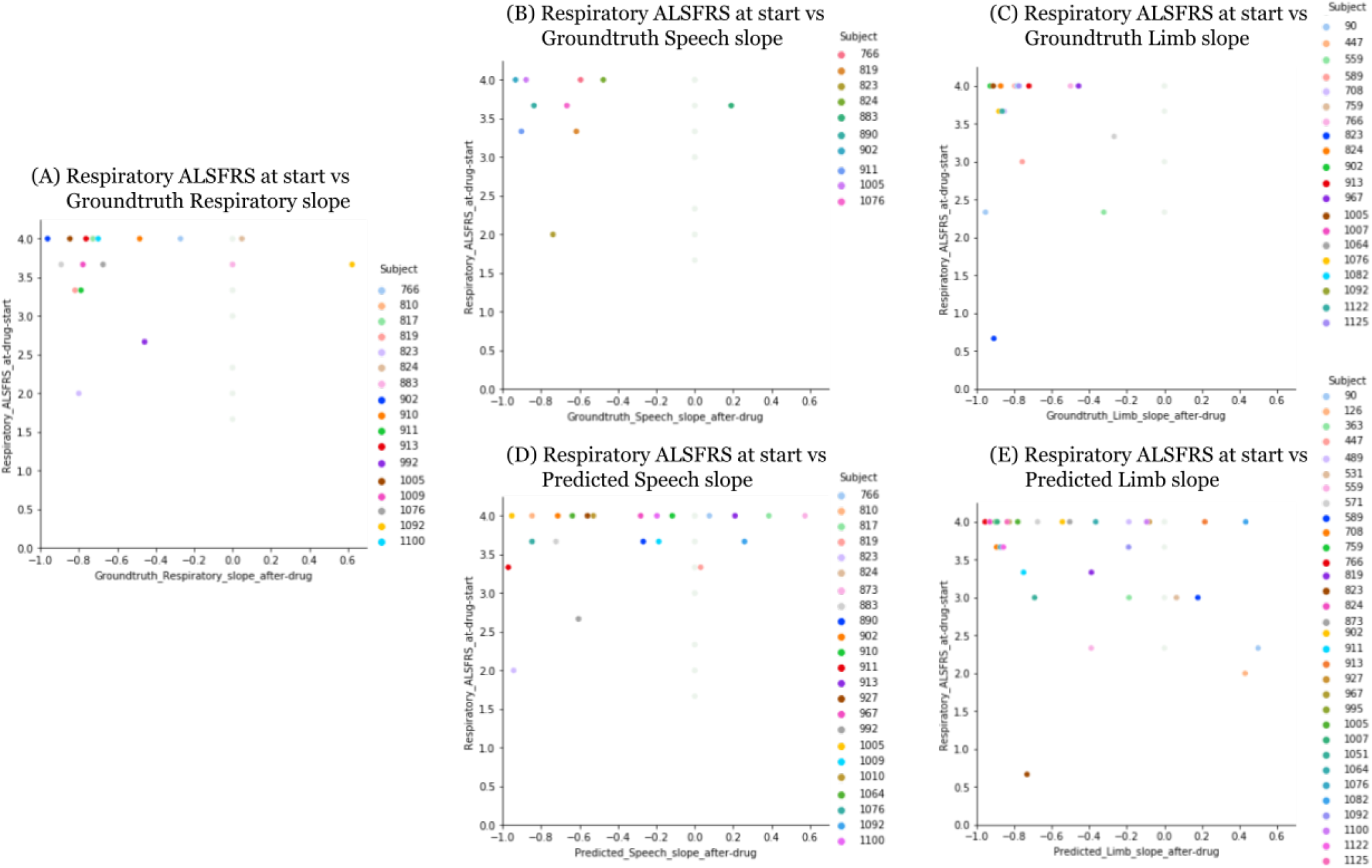
Plots comparing the average respiratory functions’ ALSFRS-R at the start of drug with the slopes (r-values) computed after starting Edavarone on the subset of 54 test participants in the Edaravone cohort. The plots depict respiratory ALSFRS-R (averaged over 3 respiratory-related functions) computed at start or pre-Edaravone and slopes (r-values) computed after (post-Edaravone) using the ALSFRS-R scores: (A) slopes from groundtruth average respiratory scores, (B) slopes from groundtruth speech scores, (C) slopes from groundtruth limb scores [averaged over 6 limb-based movements], (D) slopes from speech scores as predicted by the voice model, and (E) slopes from averaged limb-movement scores as predicted by the accelerometer model. The x-axis represents the slope (r-value) from scores after-drug (post-Edaravone), and the y-axis represents the average respiratory ALSFRS-R at or before starting Edaravone. In each plot, the IDs of participants (subjects) who had sufficient data to compute a slope after starting the drug are highlighted in color. The remaining participants whose data was insufficient to compute slope after drug are represented in a lighter color in the background (their slope is 0).

**Supplementary Figure 3 A.**
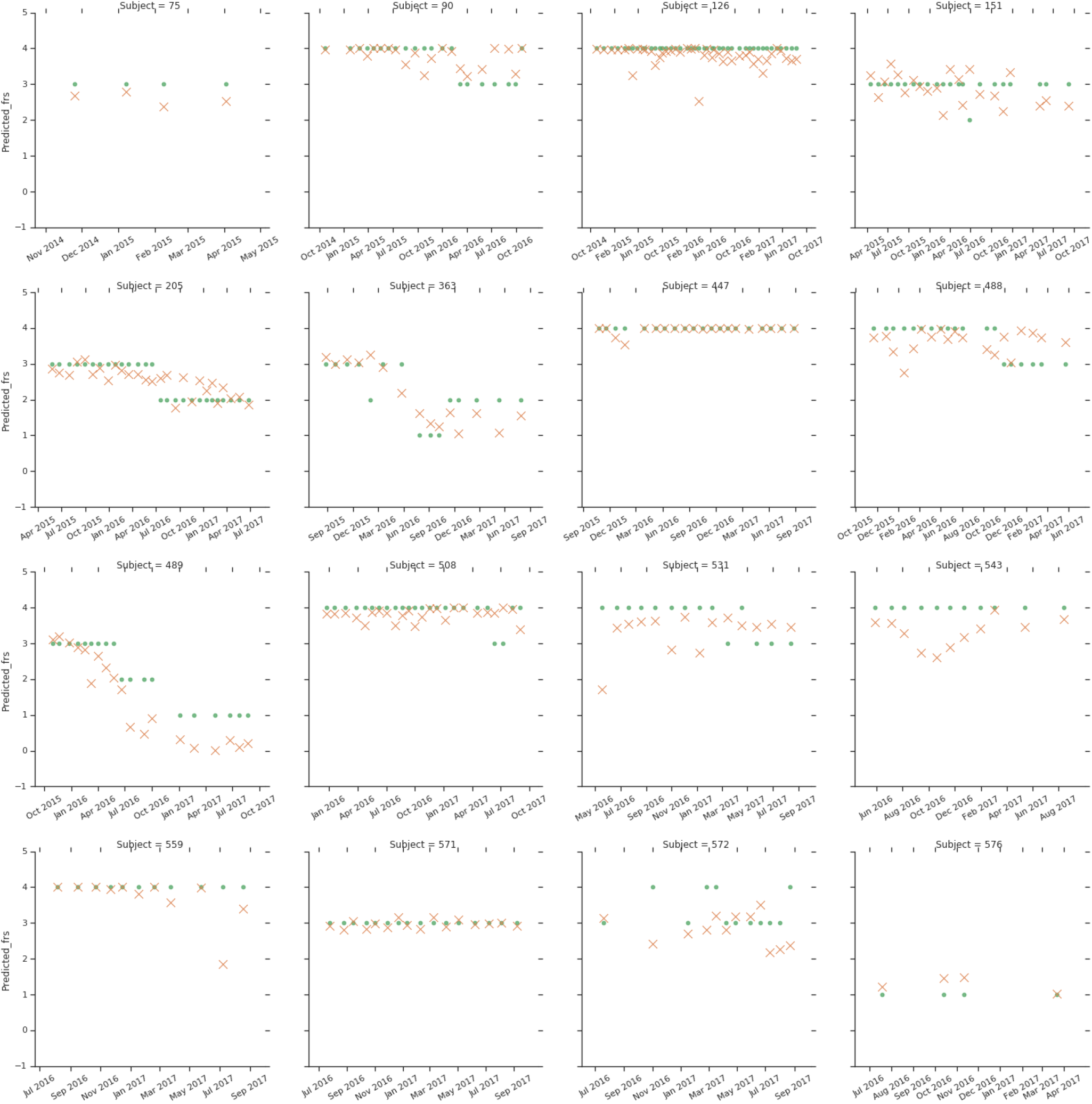
[Voice model] 1-of-3 Groundtruth and predicted labels and slopes for before and after drug for all participants in the edaravone cohort. Higher resolution is included in the supplementary materials.

**Supplementary Figure 3 B.**
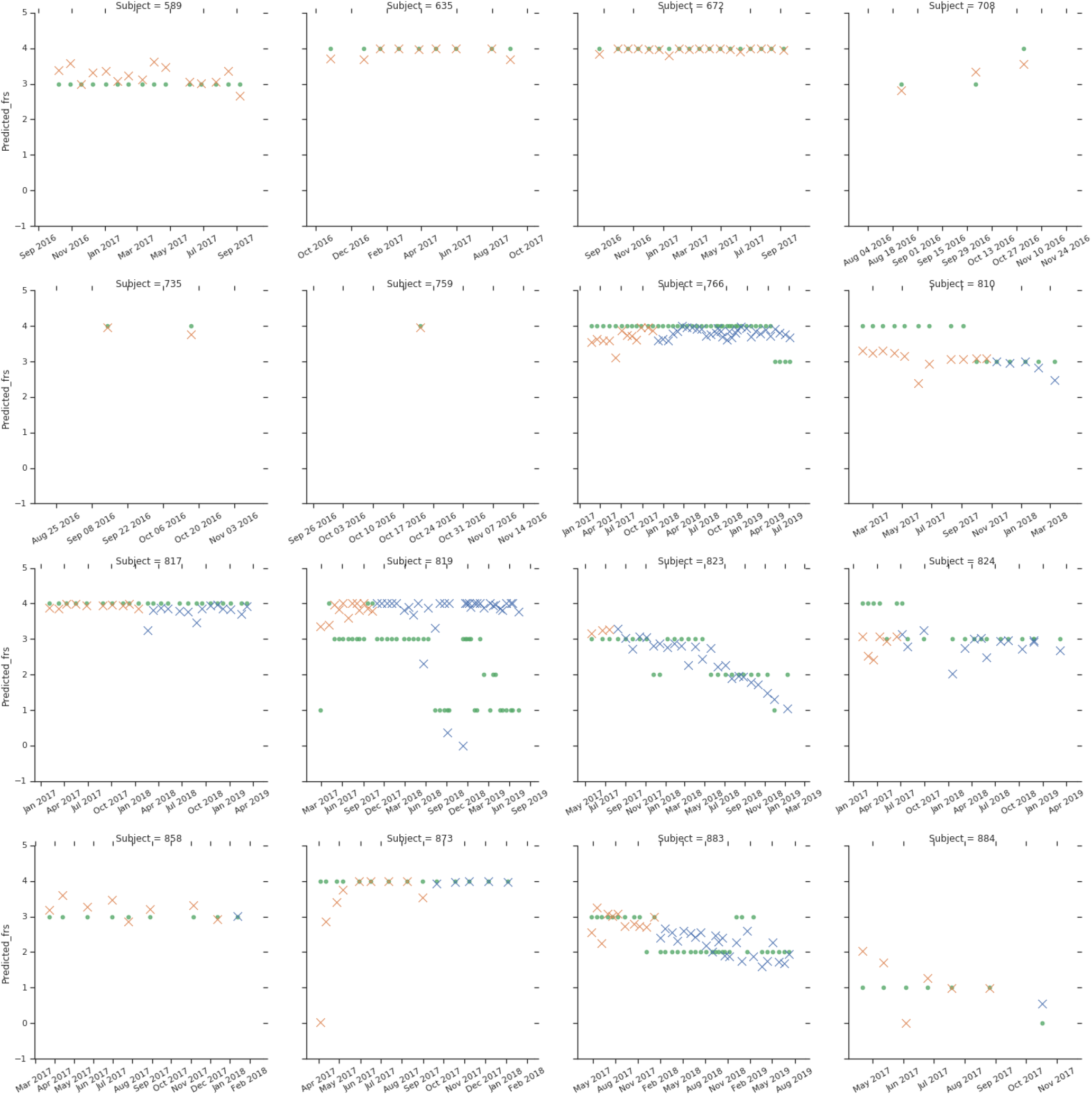
[Voice model] 2-of-3 Groundtruth and predicted labels and slopes for before and after drug for all participants in the edaravone cohort. Higher resolution is included in the supplementary materials.

**Supplementary Figure 3 C.**
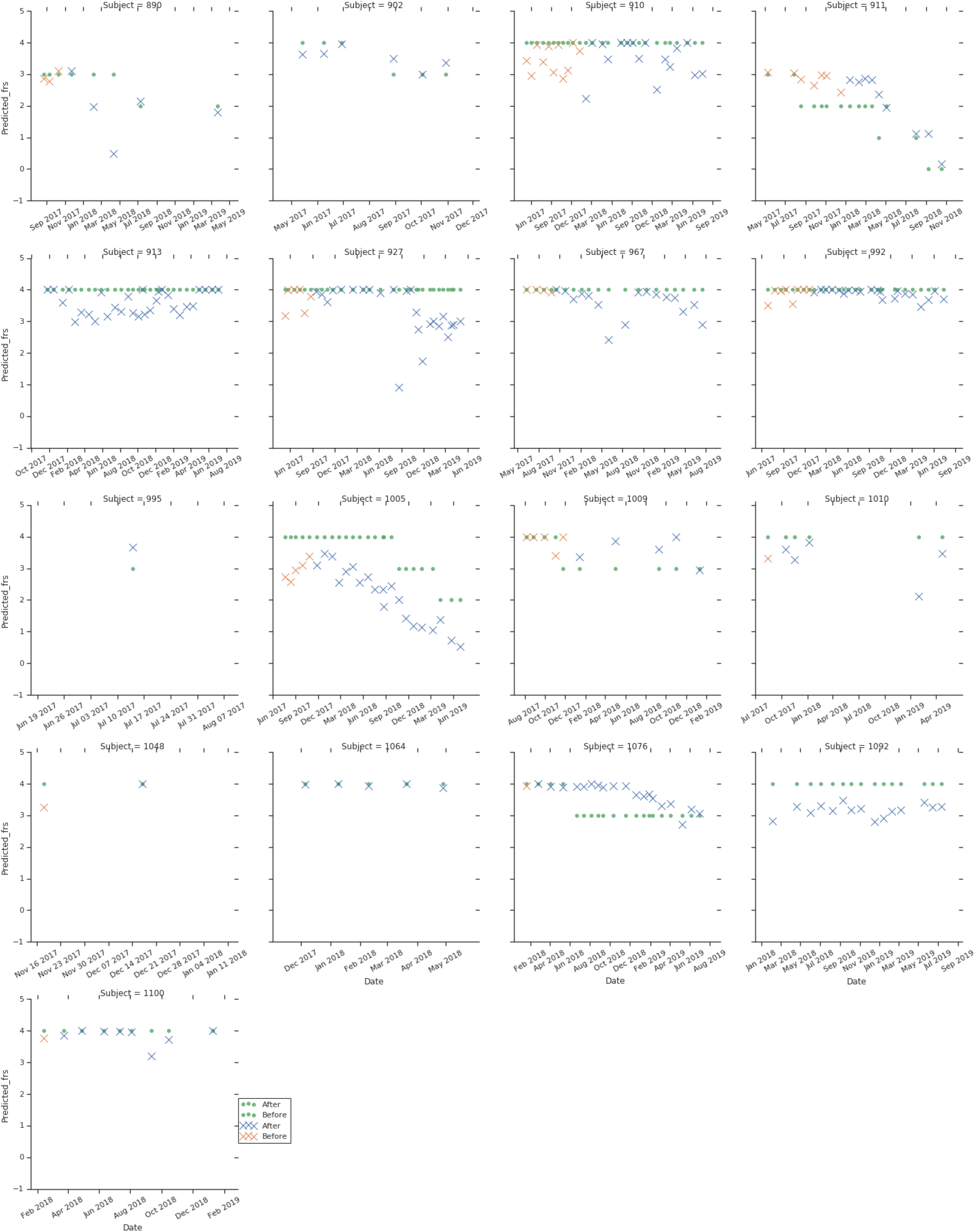
[Voice model] 3-of-3 Groundtruth and predicted labels and slopes for before and after drug for all participants in the edaravone cohort. Higher resolution is included in the supplementary materials.

**Supplementary Figure 4 A.**
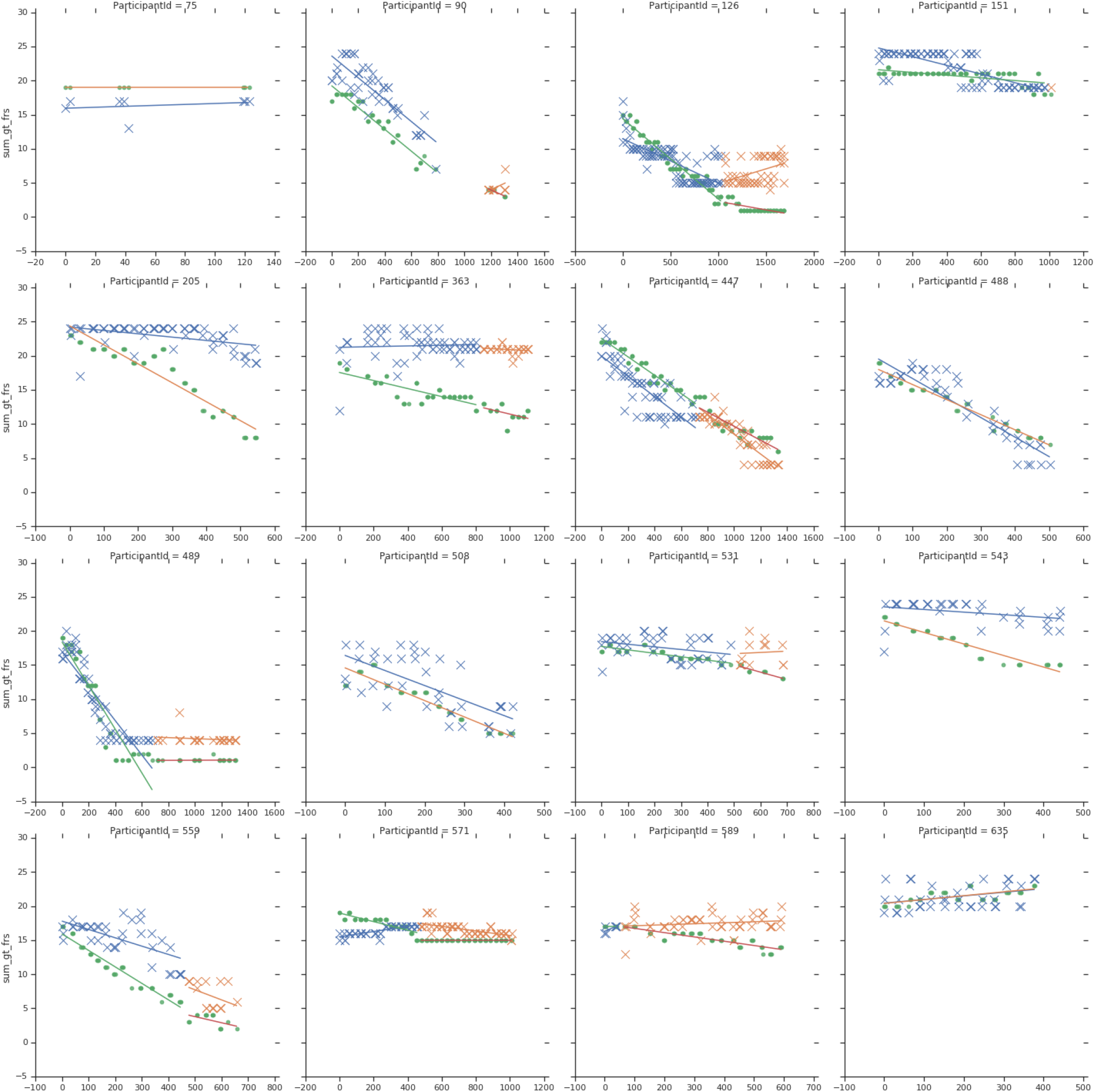
[Accelerometer model] 1-of-3 Groundtruth and predicted labels and slopes for before and after drug for all participants in the edaravone cohort. Higher resolution is included in the supplementary materials.

**Supplementary Figure 4 B.**
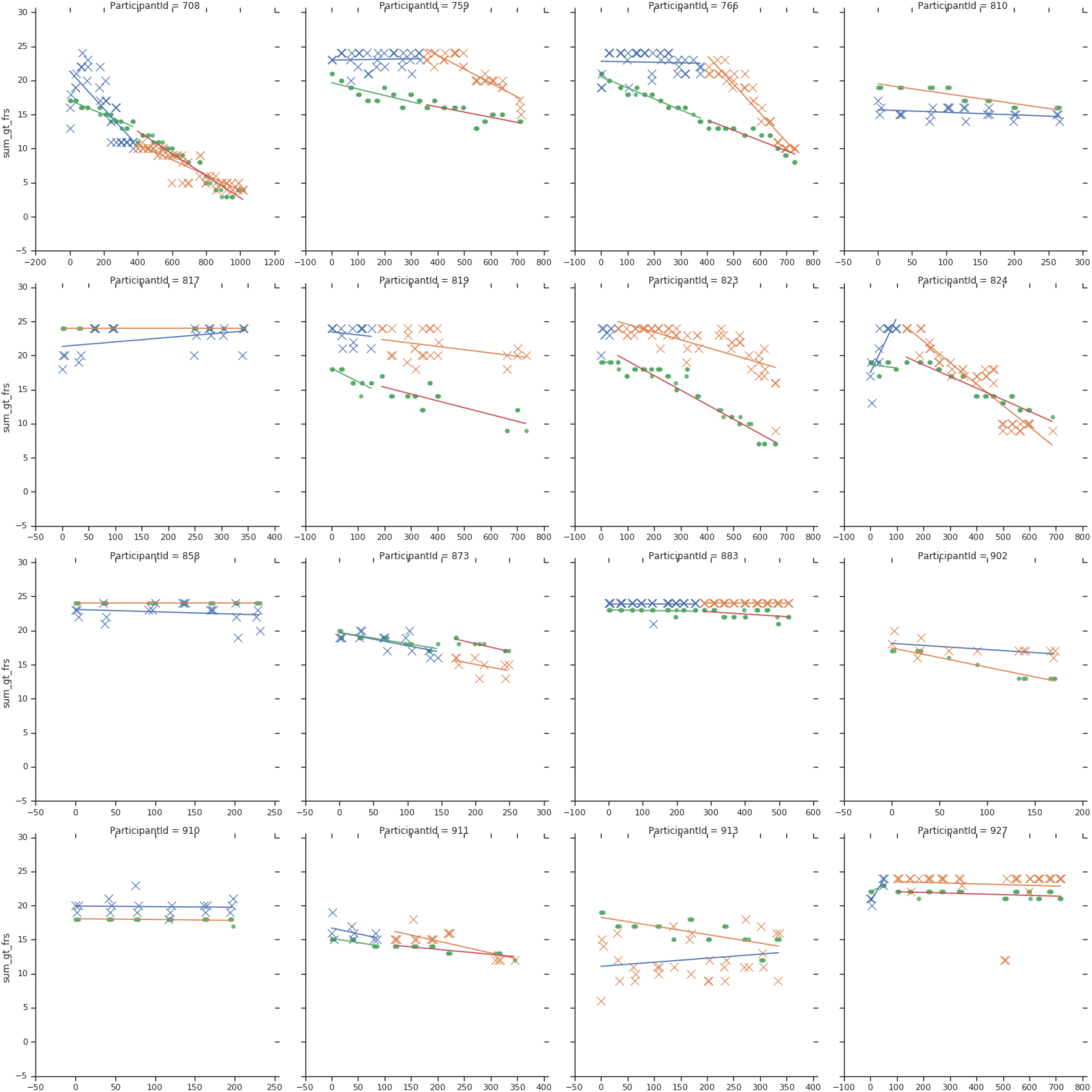
[Accelerometer model] 2-of-3 Groundtruth and predicted labels and slopes for before and after drug for all participants in the edaravone cohort. Higher resolution is included in the supplementary materials.

**Supplementary Figure 4 C.**
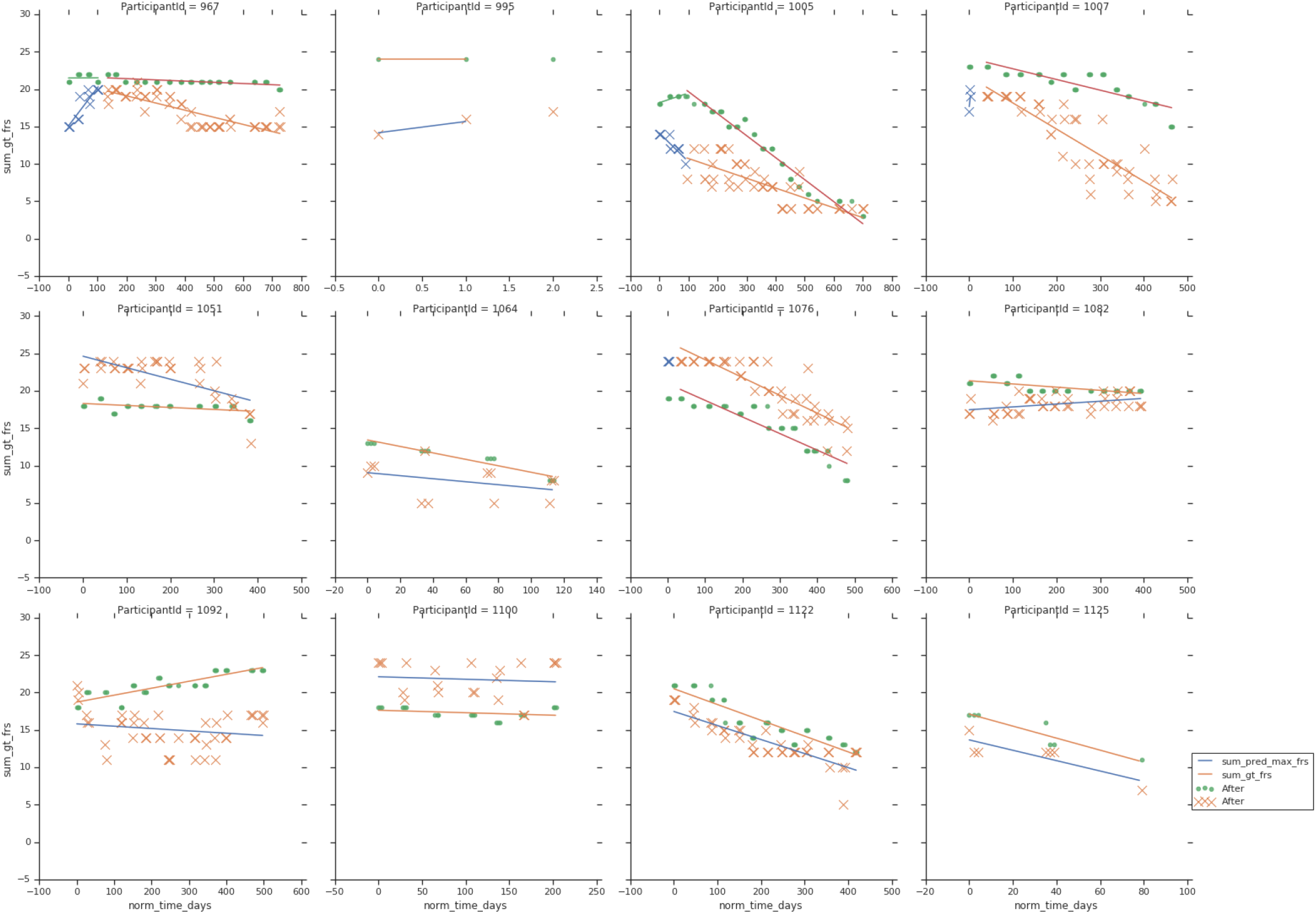
[Accelerometer model] 3-of-3 Groundtruth and predicted labels and slopes for before and after drug for all participants in the edaravone cohort. Higher resolution is included in the supplementary materials.

## Footnotes

1 To do this, ALS-TDI identified PMP participants who reported by email or through the PMP web portal that they had received treatment with edaravone. We subsequently confirmed this by telephone conversations and learned when treatment with edaravone commenced for each participant.

